# Association of Y-chromosomal gr/gr deletions with testicular germ cell tumor: whole-genome analysis of 198,306 individuals

**DOI:** 10.64898/2026.02.04.26345360

**Authors:** Subin Choi, Maria Santa Rocca, Cinzia Vinanzi, John Pluta, Zeid Kuzbari, Chey Loveday, Sophie Allen, Beth Torr, Benita Weathers, Lynn Anson-Cartwright, Darren R. Feldman, Jourik A. Gietema, Anna Gonzalez-Neira, Robert J. Hamilton, Csilla Krausz, Giovenale Moirano, Kevin T. Nead, Jérémie Nsengimana, Jenny N. Poynter, David J. Vaughn, Peter A. Kanetsky, Katherine L. Nathanson, Alberto Ferlin, Clare Turnbull, Charlie F. Rowlands, the Testicular Cancer Consortium (TECAC)

## Abstract

**Purpose:** Germline deletions affecting the Y-chromosomal gr/gr region were reported in 2005 as associated with susceptibility to testicular germ cell tumor (TGCT), a highly heritable tumor type that is the most common cancer type affecting adult men under the age of 45. Attempts to replicate this association have been equivocal, primarily due to limited power.

**Methods:** Here, we compare and validate two computational approaches to gr/gr deletion calling in high-, low– and ultra-low-coverage whole genome sequencing data, applying these to two datasets from UK Biobank and the TECAC consortium. We generate dataset-specific effect size estimates for the gr/gr deletion-TGCT association using Firth’s bias-reduced logistic regression across a total of 198,306 men of European-like ancestry (2231 with and 196,075 without TGCT).

**Results:** Upon random-effects meta-analysis of estimated effect sizes in the two datasets, we found no significant association between gr/gr deletion status and TGCT risk (combined odds ratio=1.24, 95% CI=0.74-2.07, p=0.42), nor upon stratification of seminoma and non-seminoma/mixed histological subtypes.

**Conclusion:** Our analysis suggests gr/gr deletion status alone is likely not predictive of TGCT risk in population-scale analyses of European-like individuals; however, the importance of other proposed determinants of gr/gr deletion impact, including Y-haplogroups and semen phenotype, remains unexplored at scale.

## Introduction

Testicular germ cell tumor (TGCT) comprises a group of highly heritable cancers that are the leading cancer type affecting men aged 18-44^1,2^. As a male-specific disease, variation on the Y chromosome (chrY) has naturally been posited to contribute to TGCT etiology. chrY is unique among human chromosomes in being entirely paternally inherited and displaying no meiotic recombination; it is also challenging to study due to its lack of meiotic recombination, limiting traditional linkage studies^3,4^, and an abundance of repetitive and homologous regions along its length^5–7^. One such region, the azoospermia factor c (AZFc) region, comprises 13 palindromic ampliconic sequences (Figure 1A) and is notably susceptible to the generation of copy number variants (AZFc-CNVs) arising predominantly through the process of non-allelic homologous recombination (NAHR)^8,9^. The AZFc region is one of few chrY loci with robustly demonstrated phenotypic associations, with AZFc-CNVs being the primary known genetic cause of severe spermatogenic failure (SSF)^10^. Of these, gr/gr deletions, which comprise the deletion of six of the AZFc amplicons (Figure 1B), are by a substantive margin the most frequent and most conclusively associated with SSF^11^, although evidence suggests this association may be highly dependent on genetic background^12^.

**Figure 1.**
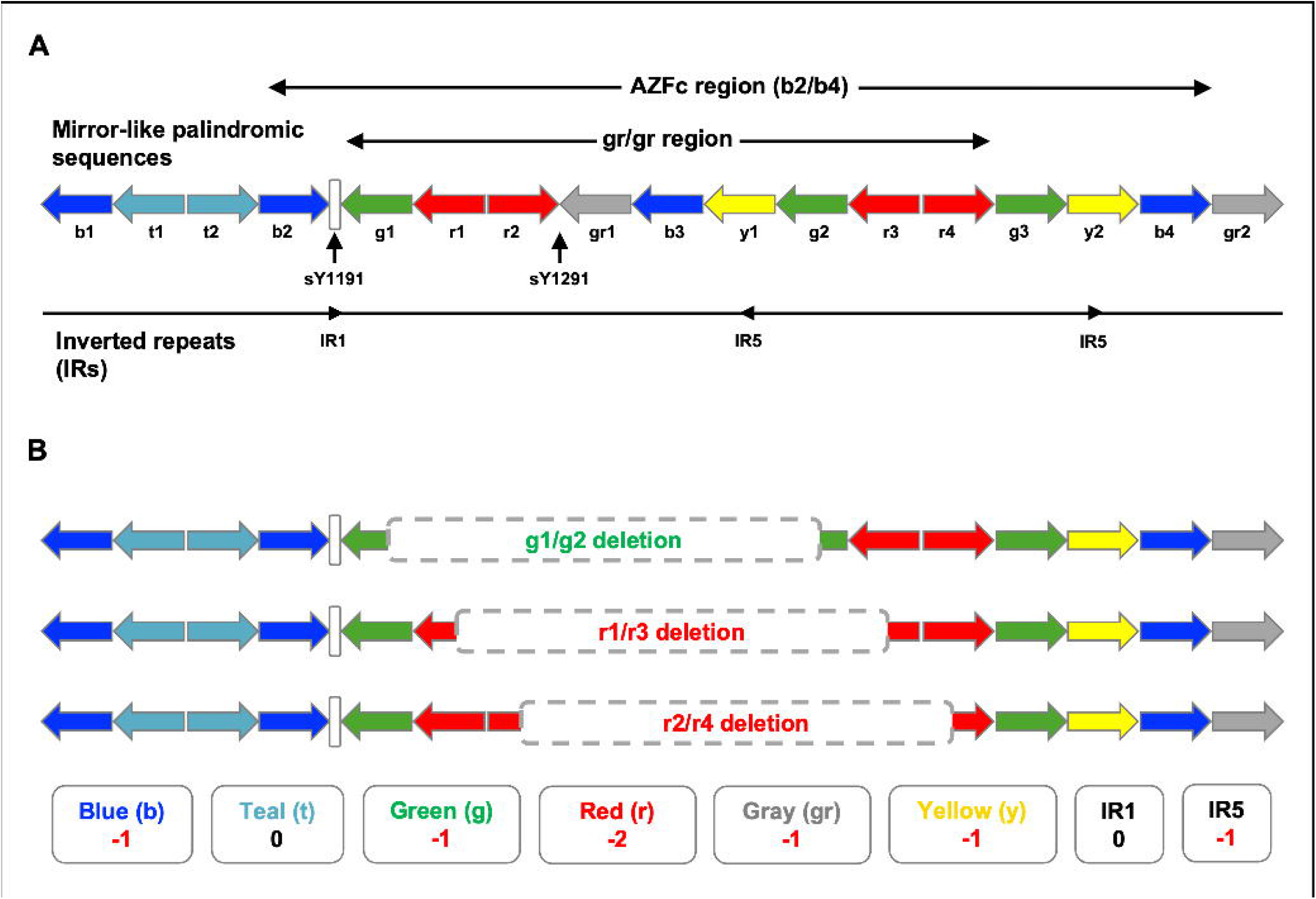
Genomic architecture of the palindromic AZFc and gr/gr regions of the human male Y-chromosome. (**A**) The human AZFc locus comprises a series of 13 palindromic ampliconic regions and lies within a larger sequence of 17 such regions, each belonging to one of six classes named for a color (top). Within the AZFc region lies the gr/gr region, spanning 9 of these ampliconic sequences from g1 to r4, inclusive. Two short sequence-tagged site markers, sY1191 and sY1291, lie just upstream of and at the r2/gr1 junction within the gr/gr region, respectively, and are used to identify gr/gr deletions in clinical diagnostics. The AZFc region further contains three inverted repeat (IR) sequences (bottom). (**B**) Three primary subtypes of gr/gr deletion have been identified (top) which differ in their precise breakpoints but result in the same overall ampliconic and IR copy number changes relative to the wild-type AZFc architecture (bottom). Adapted from Sin et al.^12^

Given the long-standing recognition of infertility as a risk factor for TGCT^13^, previous studies have investigated the possible association of gr/gr deletions with TGCT. In a 2005 study, PCR-based methods for quantification of the copy number of relevant chrY sequence-tagged site (STS) markers were used to determine the prevalence of gr/gr deletions in 1842 TGCT cases and 2599 unaffected controls, being at the time the largest study of TGCT genetic predisposition^14^. This analysis led to reporting of the gr/gr deletion as the first genetic factor conferring susceptibility to TGCT, with an estimated prevalence of 3.0% in familial TGCT cases, 2.3% in unselected TGCT cases and 1.3% in controls, yielding a logistic regression-adjusted odds ratio (OR) of 3.2 (95% CI=1.5-6.7) in familial TGCT cases and 2.1 (95% CI=1.3-3.6) in unselected TGCT cases. Subsequent molecular investigations of this association using STS-based analysis have been equivocal, primarily owing to low sample number and concomitant lack of power^15–18^.

The gr/gr deletion stood as the sole established genetic risk factor for TGCT until 2009, when the first GWAS in TGCT was published, reporting association with TGCT for five common variants^19^. Over the subsequent fifteen years, genome-wide association studies in TGCT, and subsequent meta-analyses, have proven strikingly productive, with identification of 78 loci explaining over 44% of the heritable risk of TGCT^20–27^. Notably, due to challenges in analytical methodologies, only a small proportion of GWAS studies in TGCT and other diseases have evaluated the phenotypic contribution of variation on the sex chromosomes^26^. Pathways implicated by GWAS in TGCT development include primordial germ cell (PGC) differentiation and the mitotic spindle checkpoint^21,22^. Of note, gr/gr deletions result in the deletion of two out of four members of the *DAZ* gene family (*DAZ1*, HGNC:2682; *DAZ2*, HGNC:15964; *DAZ3*, HGNC:15965; *DAZ4*, HGNC:15966) present at the AZFc locus. *DAZL* (HGNC:2685), an autosomal member of this family, is recognized as a critical determinant of PGC differentiation^28,29^. Further, *DAZL* knockouts in mice and pigs result in spontaneous gonadal teratomas^28^, providing a potential mechanistic link between GWAS-implicated pathways and gr/gr deletion biology. Via gene-level analyses of summed rare deleterious variants identified on exome sequencing, a number of genes have been proposed as TGCT susceptibility genes; as yet, replication of these findings has been inconsistent. Via candidate-based analyses, *CHEK2* (HGNC:16627) has been posited as a possible TGCT-associated gene in some studies^30,31^. A recent transcriptome-wide association study identified a further seven novel candidate TGCT-associated genes^32^.

The sparsity of chrY single nucleotide polymorphisms (SNPs) represented on typical SNP-genotyping arrays and of chrY genic regions captured by conventional exome capture kits has largely precluded informative copy number analyses using these data sources. However, over the last five years, increasing volumes of whole genome sequencing (WGS) data have been generated, much of it on population cohorts often well-characterized for their previous medical history of diseases such as cancer, and sometimes detailing follow-up via linkage to national registry data. Whilst TGCT is comparatively rare and male-specific, due to its early age of onset and excellent survival, it is commensurately well represented in such population cohorts.

The challenges of copy number calling from WGS data and discordancy between approaches have been well-documented^33,34^, further challenges on top of which are posed by chrY analysis, including that of the AZFc region, due to its repetitive nature^35^. We therefore sought to identify and build upon methodologies that had already been successfully implemented for calling of AZFc-CNVs, and to evaluate them comparatively in regard of their concordance, performance at varying sequencing depths and computational efficiency for deployment for case-control analyses across WGS datasets comprising hundreds of thousands of samples.

We present herein a comparison of two approaches for calling of gr/gr deletions through copy number analysis at AZFc ampliconic regions (Supplemental Figure 1): a bespoke AZFc-CNV caller previously published by Teitz et al. (the “Teitz-ampliconic” method)^36^ and an adaptation of the established general-use CNV caller CNVkit^37^ for calling of this region (the “CNVkit-ampliconic” method). We present cross-validation of gr/gr deletion calls made by these approaches using high-, low– and ultra-low-coverage WGS data and perform a small prospective validation of these calls against PCR-based calling, the de facto gold-standard used as a diagnostic assay. We apply the validated methodologies to quantify gr/gr deletion prevalence and association with TGCT in two large WGS datasets totaling 198,306 individuals: high-coverage data from UK Biobank (n=785 men with TGCT/195,290 men without TGCT) and ultra-low-coverage data from the Testicular Cancer Consortium (TECAC; n=1584 men with TGCT/647 men without TGCT), and present a combined effect size estimate using random-effects meta-analysis.

## Materials and Methods

### Datasets

#### 1000 Genomes Project (1KGP) dataset

Paired high-coverage (30X) and low-coverage (7.4X) whole genome sequencing data (in CRAM format) were downloaded for 1194 male samples from the 1KGP dataset via the project FTP site.

#### UK Biobank (UKB) cohort

UK Biobank (UKB) comprises genetic data from approximately 500,000 individuals recruited from the UK population aged 40-69. WGS and SNP array data (median depth ∼32.5X) were retrieved for all male participants for whom both data types were available (n=221,979) via the UKB Research Analysis Platform (UKB-RAP).

UKB samples were stratified into men with and without TGCT on the basis of both self-reported and registry-linked cancer records, resulting in a total of 846 case samples. Variant counts in cases were further stratified according to the histology of their TGCT when these data were available (seminoma vs. non-seminoma/mixed; see Supplemental Methods). Men were diagnosed with TGCT at a median age of 40, with a histology distribution of 67.6% (467/691) seminoma and 32.4% (224/691) non-seminoma/mixed among samples for which available histology information indicated either category (see Supplemental Methods). These rates may in part represent some survival bias: seminomas have better survival and are diagnosed at a later age than non-seminomas.

#### Testicular Cancer Consortium (TECAC) cohort

The TECAC cohort comprised 3342 TGCT cases and 1398 controls submitted by collaborators within the TECAC consortium from North America, Italy, the Netherlands, Spain and the UK for whom ultra-low-coverage (∼0.7X) WGS data had been generated (Supplemental Table 1). The 3342 men with TGCT had a median age of 33 at diagnosis, with a histology distribution of 52.9% (735/1390) seminoma and 47.1% (655/1390) non-seminoma/mixed. Histology data was unavailable for 1951 samples, and one sample with a histology annotation of “spermatocytic seminoma” was excluded from histological subgroup analysis (see Supplemental Methods).

### Tools for computational calling of Y chromosome AZFc CNVs

The performance of gr/gr deletion calling was compared for two bioinformatics approaches. The “Teitz-ampliconic” approach comprised a previously published bioinformatics workflow for quantification of AZFc amplicon copy numbers, for which computational AZFc-CNV calls for 1000 Genomes Project (1KGP) samples had been confirmed on validation by fluorescence *in situ* hybridization (FISH)^36^. The Teitz-ampliconic approach was implemented in Python using the publicly available GitHub package (https://github.com/lsteitz/y-amplicon-evolution). This approach required upstream conversion of WGS CRAM files to BAM format via samtools (v1.15.1)^38^.

We also developed an AZFc-specific adaptation of the general-use CNV caller CNVkit^37^, termed the “CNVkit-ampliconic” approach, as described in the Supplemental Methods and Supplemental Figure 1. In this approach, copy number changes across all six classes of AZFc amplicon and both inverted repeat (IR) sequences were quantified across bins of fixed size. Samples were then assigned either gr/gr deletion-positive or gr/gr deletion-negative status as in the existing Teitz-ampliconic pipeline.

CNVkit was implemented using a containerized docker image, and custom Python and R scripts used for downstream gr/gr deletion inference. gr/gr deletions were inferred to be present when the ampliconic copy number changes in a sample matched the established gr/gr deletion-specific signature (Supplemental Table 2). To counter batch effects and marker-specific biases, a median centering adjustment was applied to each set of ampliconic copy number estimates via both approaches (see Supplemental Methods and Supplemental Figure 2).

### Comparative validation of methodologies for calling of gr/gr deletion in lower-coverage WGS

For validation of the performance of the two methodologies at lower sequencing depths, we used the low-coverage 7.4X 1KGP dataset to generate an ultra-low-depth dataset of approximately 0.7X (a comparable depth to samples in the TECAC cohort), deploying the view command of the samtools software package^38^ (v1.15.1), setting the –s parameter to 0.1, representing a downsampling of 90%.

We were also able to perform a small-scale prospective molecular validation of the ultra-low-depth WGS calls for the TECAC samples. 79 samples from TGCT cases with sufficient remaining DNA were prospectively reanalyzed using STS-PCR, with qPCR used to confirm gr/gr deletion status in five samples. Details of the STS-PCR and qPCR experimental procedures are given in the Supplemental Methods and Supplemental Table 3.

### Case-control analysis

Twelve WGS samples from the TECAC cohort for which the Teitz-ampliconic pipeline failed to run correctly to completion were excluded from downstream analyses (Supplemental Table 4).

Autosomal SNP array data for the UKB samples were merged with autosomal WGS data for the TECAC and 1KGP datasets, retaining SNPs common to all three datasets. Principal component analysis (PCA) was conducted on the merged dataset using PLINK v2.0^39^, using the ––approx flag due to the large number of samples (Supplemental Figure 3). A supervised random forest model, implemented via the randomForest package (v4.7-1.1) in R, was trained with a 70:30 training:test split to predict the ancestral superpopulation of the pre-labelled 1KGP samples using the first ten autosomal principal components (PCs). The resulting model exhibited a classification accuracy of >0.98 (see Supplemental Methods), and was used to predict the ancestral superpopulation of each unannotated UKB and TECAC sample. Samples predicted to be of European (EUR) ancestry were retained for association analysis (Supplemental Table 4). A second round of autosomal PCA was conducted independently within the UKB and TECAC European cohorts to account for the possibility of residual population substructure.

All samples carrying non-gr/gr deletion AZFc-CNVs were excluded from association testing to avoid potential confounding in the case of association between these rarer CNV types and TGCT (Supplemental Table 4). For each of the UKB and TECAC European cohorts, association analysis between gr/gr deletions and TGCT was conducted, and individual effect size estimates combined via meta-analysis, as described below.

### Statistical analysis

All data processing and analysis was conducted using R v4.2.3 and Python v3.6 on institutional high-performance computing servers, unless otherwise stated. All metrics of association between gr/gr deletion-positive status and TGCT, and associated p-values, were generated using Firth’s bias-reduced logistic regression (implemented via the logistf package in R), given the comparative rarity of gr/gr deletion events and, in UKB, the imbalance between cases and controls^40^. The first three autosomal PCs as covariates for each dataset. Dataset-specific association effect size estimates were combined via meta-analyses under both random-effects and fixed-effect inverse variance-weighted models. Stratified association and meta-analyses were similar conducted for each TGCT histological subtype (seminoma and non-seminoma/mixed).

## Results

### Validation of computational Y chromosome copy number calling methodologies for calling gr/gr deletion in high-depth WGS data

We first sought to replicate the previously published data from Teitz et al.^36^ by running the Teitz-ampliconic pipeline on high-coverage (30X) WGS data from 1194 male samples from the 1000 Genomes Project (1KGP). gr/gr deletions were called in 47/1194 (3.94%) of 1KGP samples. The calls of samples with and without gr/gr deletions (hereafter termed gr/gr “deletion-positive” and “deletion-negative” calls) were entirely concordant with the published sample series^36^; these assignments were used as true positives and true negatives for subsequent validation using 1KGP data.

Notably, this methodology had an average runtime of 2 hours and storage of 21 GB per sample, rendering it unfeasible for deployment at scale on WGS data for the approximately 200,000 male samples in UKB. We thus sought to adapt the established and computationally efficient CNV caller CNVkit^37^ for gr/gr deletion calling (Supplemental Figure 1; Methods).

Calls from this “CNVkit-ampliconic” approach entirely recapitulated those from the Teitz-ampliconic approach (47/47 gr/gr deletion-positive and 1147/1147 gr/gr deletion-negative). The CNVkit-ampliconic approach required only 14 minutes and 7 GB storage per sample, rendering it feasible for deployment for high-coverage, large-scale population sequencing datasets.

### Validation of computational Y chromosome copy number calling tools for calling gr/gr deletion in lower-coverage WGS data

The largest number of men with TGCT available to our analysis was contributed by the ultra-low-coverage (median 0.7X) TECAC WGS dataset. We therefore sought to establish the performance of our copy number calling tools on low-coverage WGS data. We firstly explored calling in this low-coverage dataset by applying both calling methodologies to the existing paired low-coverage (7.4X) WGS data for the 1194 1KGP samples, as well as to simulated ultra-low-coverage (0.7X) WGS data (see Methods).

The Teitz-ampliconic approach maintained excellent sensitivity for detection of gr/gr deletion (100% and 97.9%) at low (7.4x) and ultra-low (0.7x) coverage, respectively, with a false positive rate (FPR) of 0% at both lower depths (Table 1). The CNVkit-ampliconic caller exhibited comparable performance at low coverage, retaining a sensitivity of 100% and with a low FPR of 2.1%. Sensitivity for the CNVkit-ampliconic caller decreased modestly for ultra-low-coverage WGS to 93.6% with an FPR of 2.2%.

**Table 1.** Quantification of gr/gr deletion calling performance of two bioinformatics approaches in low (7.4X) and simulated ultra-low (0.7X) 1KGP WGS data. The sensitivity and specificity of two bioinformatics approaches for calling of gr/gr deletions were compared, using as the positive gr/gr deletion truthset the 47 consensus gr/gr deletion calls identified via both the Teitz-ampliconic and CNVkit-ampliconic approaches when applied to high-coverage (30X) WGS data from 1194 1KGP samples.

|  |  | Prediction in lower-coverage WGS data |  |  |  |  |  |  |  |
| --- | --- | --- | --- | --- | --- | --- | --- | --- | --- |
|  |  | Low-coverage (7.4X) |  |  |  | Simulated ultra-low-coverage (0.7X) |  |  |  |
|  |  | Teitz-ampliconic |  | CNVkit-ampliconic |  | Teitz-ampliconic |  | CNVkit-ampliconic |  |
|  |  | gr/gr deletion-positive | gr/gr deletion-negative | gr/gr deletion-positive | gr/gr deletion-negative | gr/gr deletion-positive | gr/gr deletion-negative | gr/gr deletion-positive | gr/gr deletion-negative |
| Prediction in high-coverage (30X) WGS data | gr/gr deletion-positive | 47 | 0 | 47 | 0 | 46 | 1 | 44 | 3 |
|  | gr/gr deletion-negative | 0 | 1147 | 1 | 1146 | 0 | 1147 | 1 | 1146 |
|  | Sensitivity | 1 |  | 1 |  | 0.979 |  | 0.936 |  |
|  | Specificity | 1 |  | 0.999 |  | 1 |  | 0.999 |  |
|  | False positive rate | 0 |  | 0.021 |  | 0 |  | 0.022 |  |

We also were able to conduct a small-scale molecular validation of the performance of the Teitz-ampliconic and CNVkit-ampliconic tools for calling gr/gr deletions in ultra-low-coverage WGS by examining concordance with prospective analysis of samples from 79 TGCT cases with available DNA using PCR (STS-PCR for all 79 samples, with more precise confirmation via qPCR for five samples; Supplemental Figure 4). gr/gr deletion calls from the ultra-low-coverage WGS for each of the Teitz-ampliconic and CNVkit-ampliconic tools were fully concordant with the calls from PCR analysis, with 76/76 samples concordantly assigned as gr/gr deletion-negative and 3/3 samples concordantly assigned as gr/gr deletion-positive.

Notably, one sample called as gr/gr deletion-negative using qPCR (the more sensitive methodology) had initially been called as gr/gr deletion-positive using STS-PCR; the qPCR gr/gr deletion-negative call was consistent with both computational approaches. This sample was noted to be older and of lower quality DNA (Supplemental Figure 4).

### Inference of gr/gr deletion hemizygote status among TECAC and UKB participants

Using PCA, a total of 4113 individuals of European-like ancestry were identified in the TECAC dataset, and 207,835 in UKB (Supplemental Figure 3). Based on its superior sensitivity at ultra-low coverage, we called gr/gr deletions using the Teitz-ampliconic approach in these TECAC samples (median WGS depth 0.7X); for the UKB samples (median WGS depth 32.5X), the CNVkit-ampliconic workflow was applied due to its computational efficiency at high-coverage. We observed a gr/gr deletion hemizygote frequency among the European-like TECAC samples of 1.22% (50/4113) and among the European-like UKB samples of 1.42% (2943/207,835).

After excluding samples predicted to harbor other (non-gr/gr deletion) AZFc-CNV events (see Methods), a total of 2231 TECAC (1584 men with TGCT, 647 without) and 196,075 UKB (785 men with TGCT, 195,290 without) samples were retained for downstream association analysis.

### Case-control analysis for association of Y-chromosome AZFc gr/gr deletion with TGCT

Among these 2231 European-like TECAC samples, the frequency of gr/gr deletion in men with TGCT was 2.40% (38/1584) and in the men without TGCT was 1.85% (12/647). In unadjusted association analysis, this yielded an OR of 1.30 (95% CI=0.68-2.51, χ^2^ p=0.53). Undertaking logistic regression, incorporating the top three autosomal principal components of the TECAC dataset, yielded an adjusted OR of 1.69 (95% CI=0.86-3.33, p=0.12; Table 2; Figure 2).

**Figure 2.**
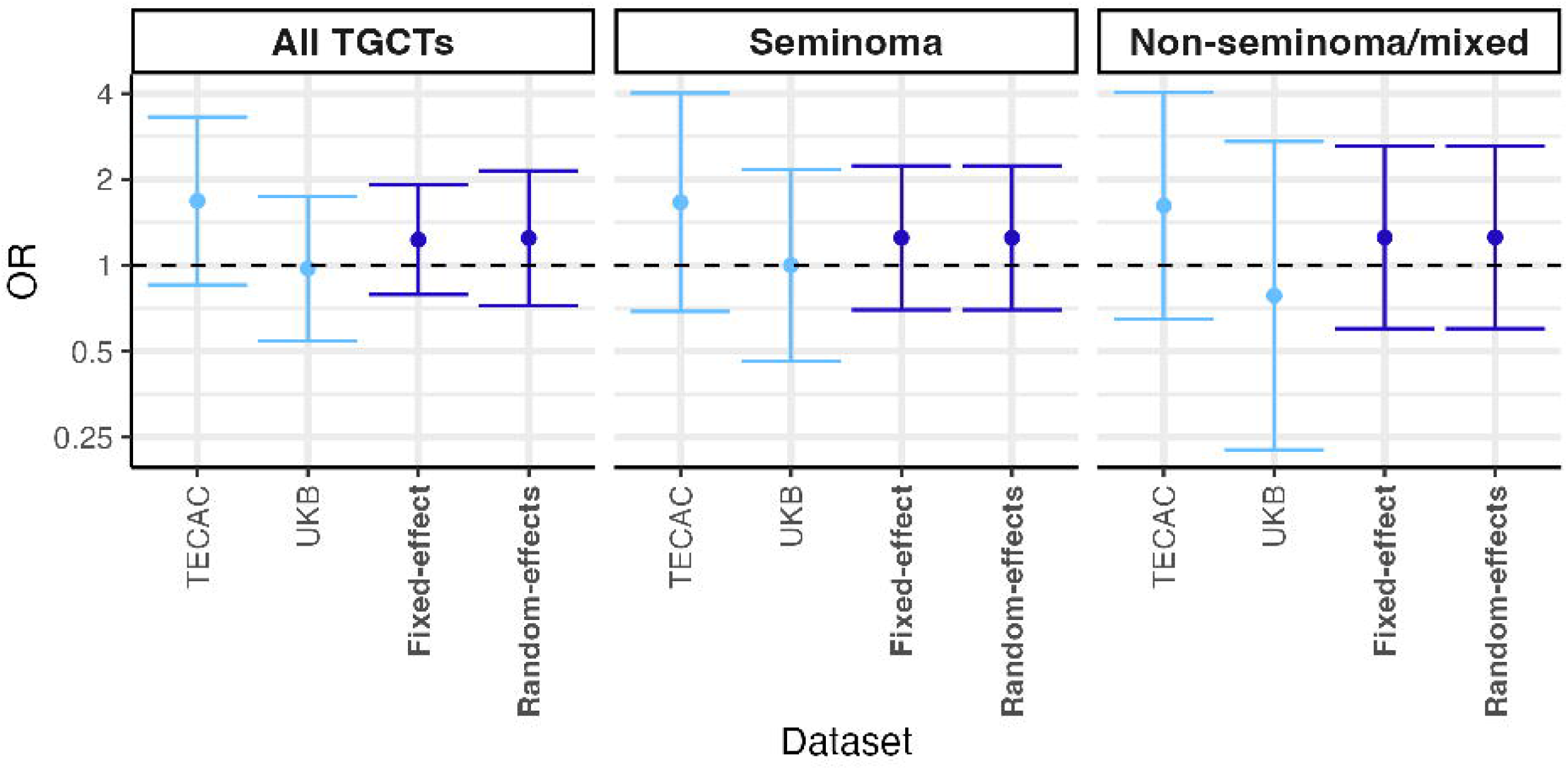
Estimated effect sizes of association between gr/gr deletions and TGCT risk, stratified by histological subtype. Shown are estimated odds ratios, as given in Table 2 and Supplemental Table 5, in each of the TECAC and UKB datasets (calculated via Firth’s bias-reduced logistic regression, adjusting for the first three autosomal PCs within each dataset), alongside two meta-analytical approaches, fixed-effect and random-effects inverse variance-weighted meta-analysis (dark blue). No evidence of association was observed in any constituent dataset or using either meta-analytical methodology. Error bars represent the 95% confidence intervals around the point estimate of the odds ratio, calculated based on the regression-calculated standard error. OR, odds ratio; TGCT, testicular germ cell tumor.

**Table 2.**
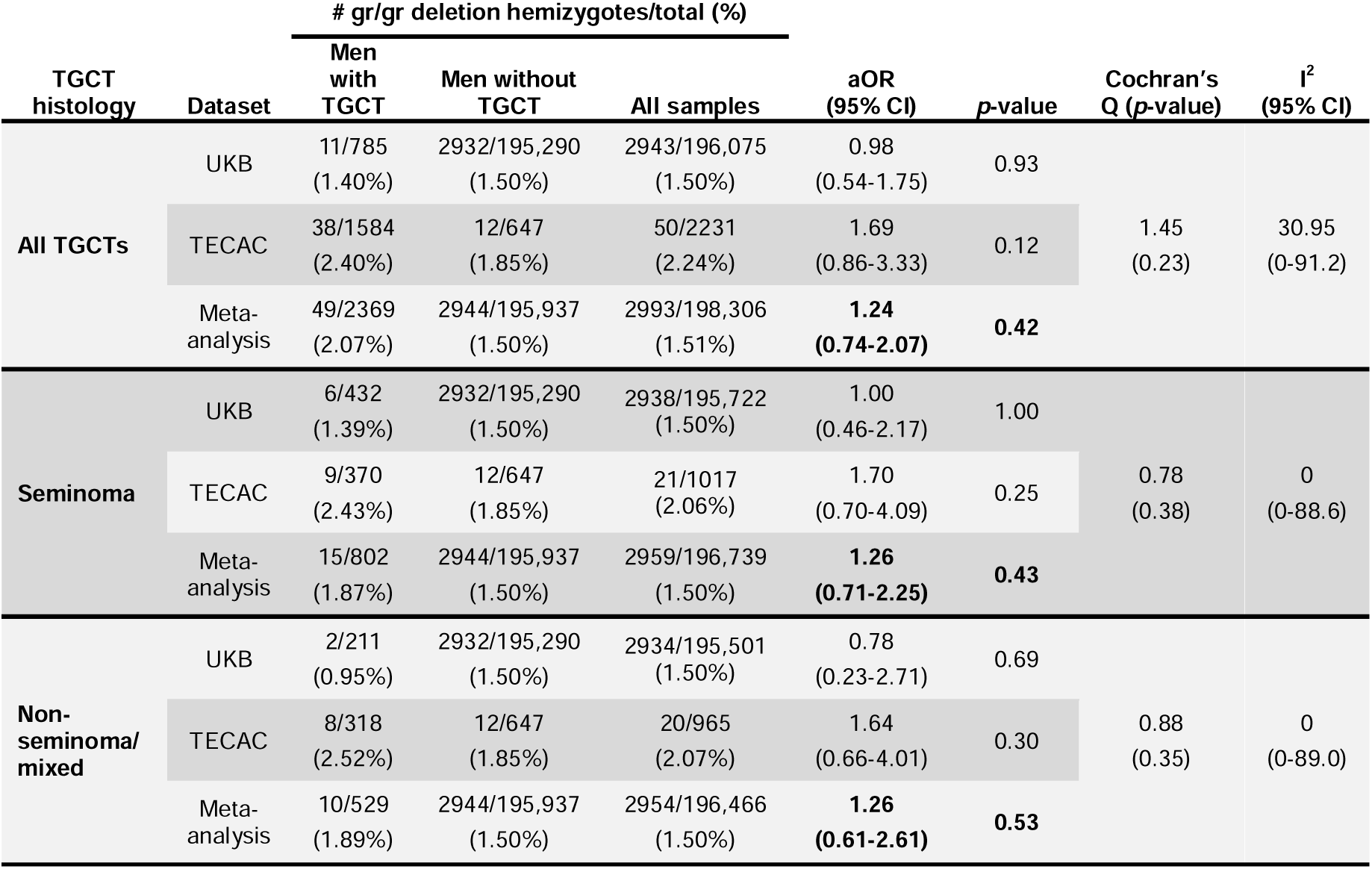
Estimated effect sizes for association between gr/gr deletions and TGCT. Odds ratios for the association between gr/gr deletion hemizygote status and TGCT were estimated using Firth’s bias-reduced logistic regression in each of the UKB and TECAC datasets. A random-effects meta-analysis with inverse-variance weighting was used to produce combined estimates of effect size. Estimates were generated for both men with TGCT overall and for each of the histological subtypes (seminoma or non-seminoma/mixed) where histology was available for samples. Two heterogeneity metrics, Cochran’s Q (alongside its *p*-value) and I^2^ are additionally presented, with I^2^ broadly interpretable as follows: 0-40% = may be unimportant; 30-60% = may represent moderate heterogeneity; 50-90% = may represent substantial heterogeneity; 75-100% = considerable heterogeneity. aOR, odds ratio adjusted using Firth’s bias-reduced logistic regression; CI, confidence interval; TGCT, testicular germ cell tumor.

Among these 196,075 UKB European-like samples, the gr/gr deletion was detected at a frequency of 1.40% (11/785) in men with TGCT and 1.50% (2932/195,290) in men without TGCT. Unadjusted calculation of effect size yielded an OR of 0.93 (95% CI=0.51-1.69, χ^2^ p=0.93). Undertaking logistic regression with inclusion of the top three autosomal PCs as covariates yielded an adjusted OR of 0.98 (95% CI=0.54-1.75, p=0.93; Table 2; Figure 2).

Given some indication of possible heterogeneity based on the I^2^ estimates (Table 2), we combined the UKB and TECAC OR estimates using random-effects inverse variance-weighted meta-analysis, which yielded a non-significant combined odds ratio for all TGCT histologies of 1.24 (95% CI=0.74-2.07, p=0.42). There was similarly no evidence of association of association on restriction of meta-analysis by histological subtype, nor upon repeating meta-analysis under a fixed-effect model (Figure 2; Supplemental Table 5).

## Discussion

We present a comparative evaluation and validation of two computational approaches for gr/gr deletion calling: an amplicon-based method described by Teitz et al.^36^, whereby CNV calling is conducted across pre-defined AZFc amplicons of variable length, and a more computationally efficient CNVkit-based approach described herein, in which AZFc amplicon copy number is quantified across fixed-size bins. Although extendable to rarer AZF-CNVs, larger datasets are required to sufficiently empower association analyses for, and validate calling of, such events. Focusing specifically on calling of gr/gr deletions, the most frequent AZFc-CNV, we demonstrate calls from both methodologies to be entirely concordant in high-depth WGS from 1194 samples in 1KGP. The sensitivity of both approaches remains 100% at 7.4X, with modest decreases to 97.9% and 93.6% at 0.7X for the Teitz-ampliconic and CNVkit-ampliconic approach, respectively. Specificity was near-perfect for both methodologies across all depths. The high computational requirements of the Teitz-ampliconic approach are mitigated in ultra-low-coverage WGS data, but render it unsuitable for application to large high-coverage sample series, such as UKB. Given its comparable sensitivity to the Teitz-ampliconic approach using high-depth WGS data, we proceeded with the computationally viable CNVkit-ampliconic methodology for the UKB analysis.

Despite good power, restriction to samples of European ancestry and additional adjustment for residual autosomal population substructure, there was no statistically significant evidence of association between gr/gr deletions and TGCT in either UKB (785 men with TGCT; 195,290 men without TGCT; OR=0.98; 95% CI=0.54-1.75; p=0.93), nor in the TECAC dataset (1584 men with TGCT; 647 men without TGCT; OR=1.69; 95% CI=0.86-3.33; p=0.12). Association was also non-significant upon fixed-effect meta-analysis (OR=1.24; 95% CI=0.74-2.07; p=0.42).

In the original 2005 report^14^, gr/gr deletions were reported to confer a three-fold increase in TGCT risk among cases with a family history of TGCT (OR=3.2; 95% CI=1.5-6.7; p=0.0027), and a two-fold increase among cases without a family history (OR=2.1; 95% CI=1.3-3.6; p=0.005; Supplemental Table 6). The association was reported to be stronger for (and significant solely for) seminoma (OR=3.0; 95% CI=1.6-5.4; p=0.0004) than non-seminoma (OR=1.5; 95% CI=0.72-3.0; p=0.29). The 1842 (including 1377 sporadic and 431 familial) cases were assembled from geographically diverse collections of modest-sized case-only series (n=1464 cases), contemporaneously recruited cases and controls (n=266 cases and n=953 controls) and five stand-alone control series (n=1668; Supplemental Table 7). STS-PCR-based gr/gr deletion genotyping was performed at two centers. Given the small proportion of contemporaneously ascertained cases and controls, consequent potential for stratification, and rarity of the gr/gr deletion, the authors highlighted the value of replication analyses.

Although other studies in the intervening period have examined the frequency of gr/gr deletions in TGCT cases and its association with disease, these follow-up studies have been much smaller and thus underpowered to robustly investigate the reported association. In 2007, Ferlin et al. identified none of the AZFc-CNVs of interest in 118 TGCT cases or 93 controls recruited in the healthcare setting in North Italy^15^. In 2019, Moreno-Mendoza et al. reported a gr/gr deletion frequency of 2.8% (14/497) in TGCT cases and 2.0% (40/2030) in controls recruited in Italy and Spain, yielding an OR of 1.44 (95% CI=0.72-2.73; p=0.24)^17^.

Thus, the current analysis represents the first study sufficiently powered to evaluate the gr/gr deletion-TGCT association reported twenty years ago.

### Limitations

As is common in analyses of CNVs in large-scale NGS datasets, the validation of calling approaches, and methodology used to define “truthsets”, was complex and challenging. Our focus on a single CNV somewhat simplified the validation. Our initial Teitz-ampliconic analysis recapitulated gr/gr deletion calls made in high-depth 1KGP WGS (and validated by FISH) in the original publication^36^. Concordance between these calls and those made by the CNVkit-ampliconic approach gave us a reasonable “ground truth” by which to establish a set of gr/gr deletion-positive (n=47) and gr/gr deletion-negative (n=1147) 1KGP samples. These were then used for validation of calling at lower depths. An optimal validation would require use of an orthogonal molecular methodology, such as PCR, to corroborate directly and comprehensively these 1KGP calls. Nonetheless, a modest-sized prospective molecular validation confirmed full concordance between gr/gr deletions called on ultra-low depth WGS by both computational methods and wet-lab PCR methods, although availability of DNA limited the size of this laboratory-based validation experiment.

For one sample noted to be older and thus of potentially lower quality, STS-PCR analysis initially called gr/gr deletion-positive status: gr/gr deletion-negative status was only called on subsequent evaluation by qPCR. As the most widely used and longstanding diagnostic methodology, STS-PCR would likely be widely regarded as the gold-standard approach for gr/gr deletion genotyping. This discordant gr/gr deletion call by STS-PCR against the more sensitive methodology of qPCR highlights the fallibility of some “established” laboratory approaches.

The TECAC series contains a limited number of men without TGCT; within UKB there is a large number of men without TGCT, but TGCT frequency is constrained by its rarity in a population cohort. The power of the overall analysis is necessarily reduced by this case-control imbalance between the two series, although we attempt to mitigate this reduction through implementation of Firth’s bias-reduced logistic regression.

Two potential confounders were not included in this analysis: Firstly, due to lack of sperm phenotype data in either dataset, we could not assess its role as a possible confounder, noting a report by Moreno-Mendoza et al. of 497 men with TGCT, in which significantly increased TGCT risk was reported only when controls were normozoospermic^17^. We were also unable to characterize Y-haplogroups (Y-Hgs) for samples due to the computational requirements of doing so across the large sample series; gr/gr deletion prevalence is known to vary across Y-Hgs, although no Y-Hg has been consistently reported as associated with TGCT. Some residual Y-chromosomal population substructure may therefore remain that was not accounted for by autosomal PCA. We additionally lacked information on family history of TGCT in either cohort, noting that gr/gr deletion prevalence was reported to be higher in familial than sporadic cases in the original 2005 association analysis^14^.

Thus, we present the first large-scale analysis of the association between gr/gr deletions and TGCT since the original 2005 report of the gr/gr deletion as the first genetic susceptibility factor for TGCT identified from large-scale analysis. Our analyses using computational gr/gr deletion calling in 2369 men with TGCT and 195,937 men without TGCT does not support significant association with unselected TGCT in men of genomically-defined European ancestry. Notably, the point estimate of association from combined meta-analysis, OR=1.24 (95% CI=0.74-2.07; p=0.42), is not encompassed within the 95% confidence intervals of the point estimate of effect size from the 2005 report of 2.1 (95% CI=1.3-3.6), based on 1842 TGCT cases and 2599 controls of self-reported white ancestry.

The compounded rarity of both TGCT and gr/gr deletion events limits our power to negate association between the two. Nevertheless, our analyses do not support gr/gr deletion hemizygote status alone being predictive of increased TGCT risk for population-ascertained cases.

## Data availability

Users may download 1KGP data via the FTP page at https://ftp.1000genomes.ebi.ac.uk/vol1/ftp/. Access to UKB data is granted to eligible researchers subject to project approval by the UKB data access team. TECAC data is available upon application to the TECAC consortium. Code for the CNVkit-ampliconic pipeline has been made available via a public GitHub repository at https://github.com/instituteofcancerresearch/CNVkit-AZFcAmpliconic.git.

## Acknowledgements

This research has been conducted using the UK Biobank Resource under Application Number 76689. We thank all patients and their clinicians for participation in this study. C.K. would like to thank Dr M. Vannucci and Dr V. Rosta from the University Hospital of Careggi and Dr D. Moreno Mendoza from the Fundacio Puigvert. D.J.V. and K.L.N. would like to thank Linda Jacobs and Donna Pucci for their contributions to participant recruitment and the study participants from the University of Pennsylvania. J.N. would like to thank Louise Parkinson for coordinating recruitment.

## Author contributions

Conceptualization: S.C., C.F.R, C.T., K.L.N., P.A.K. and C.L; Data curation: S.C., J.P., B.W., K.L.N.; Formal analysis: S.C.; Funding acquisition: C.T., K.L.N., P.A.K., A.F.; Investigation: S.C., M.S.R., C.V.; Methodology: S.C., C.T., C.L., C.F.R.; Project administration: B.W., J.P., K.L.N., P.A.K.; Resources: B.W., A.F., M.S.R.; Software: S.C.; Supervision: C.T., C.F.R., C.L.; Visualization: S.C., C.F.R., M.S.R.; Validation: S.C., M.S.R., C.V.; Writing – original draft: S.C., C.T., C.F.R., Writing – review & editing: all authors.

## Ethics declaration

Participants in the *Post genome wide association studies in Testicular Germ Cell Tumor* conducted by the **Te**sticular **Ca**ncer **C**onsortium (TECAC) were recruited with informed consent following review by the respective ethics committees of each participating site. The University of Pennsylvania (Upenn) is the coordinating site for the TECAC, thus there is separate approval by the Upenn Institutional Review Board (IRB) #7 to oversee all activities of the research collaboration, including sample collection, upload of phenotype data to the NCI research portal, and material and data transfer agreements with each site for these operations. The study was reviewed and approved by the IRB and conducted in accordance with University of Pennsylvania human subjects requirements and applicable federal regulations, and conducted in accordance with the principles of the Declaration of Helsinki.

