## Supplemental Material for "Association of Y-chromosomal gr/gr deletions with testicular germ cell tumor: whole-genome analysis of 198,306 individuals"

### Supplemental Methods

#### Comparison of computational methodologies for gr/gr deletion calling and development of a high-throughput calling approach

The analyses in this manuscript utilized two methodologies for gr/gr deletion calling; a summary of the key differences between these approaches are summarized below and in Supplemental Figure 1.

##### Calling of gr/gr deletions using the Teitz et al. ampliconic copy number calling methodology

The 2018 publication by Teitz et al. presented a copy number calling pipeline tailored specifically to analysis of CNVs affecting the AZFc region (AZFc-CNVs) of the Y-chromosome, hereafter termed the Teitz-ampliconic approach^1^. In this methodology, baseline haploid coverage is calibrated using the read depth across a 1 Mbp single-copy region of the Y chromosome (chrY). Copy number is then calculated across the coordinates of AZFc ampliconic regions of interest. This approach allows variability in the length of target regions and ensures the copy number is only quantified across these ampliconic regions, without the inclusion of any adjacent off-target sequence.

The Teitz-ampliconic approach leverages existing knowledge of the architecture of the AZFc region, which consists of a series of ampliconic units believed to result from ancestral duplication events and thereby sharing substantial sequence homology. AZFc-CNVs are believed to predominantly arise through non-allelic homologous recombination (NAHR).

In their publication, Teitz et al. used the molecular principles of NAHR to computationally model all possible AZFc-CNVs resulting from 1-, 2- or 3-step NAHR events^1^. They identified 14 distinct classes of rearrangement, each characterized by a unique signature of amplicon copy number changes. It was therefore possible to map any amplicon copy number changes observed in WGS data to corresponding AZFc-CNVs. The amplicon copy number changes corresponding to specific classes of AZFc-CNV are shown in Supplemental Table 2, with class v1 comprising the gr/gr deletions investigated in this study.

Although our analysis focused only on the gr/gr deletion as a previously suggested risk factor for TGCT, our studies indicate that these methods can be used to evaluate AZFc copy number changes in association with other phenotypes.

The Teitz-ampliconic pipeline is not readily able to take in the CRAM file format, which is currently the primary file format in which WGS data is stored. Further, as the pipeline was primarily designed for the specific analysis including within its corresponding publication^1^, cleaning and adaptation of its Python scripts was necessary for the analyses presented here.

##### Calling of gr/gr deletions using the CNVkit-ampliconic approach

CNVkit is an established general-purpose bioinformatics tool for the calling of copy number of regions of interest from high-throughput sequencing data^2^. The user first specifies on-target and off-target (also known as antitarget) regions: the former consists of the regions in which copy number is to be called, with the latter representing the regions used to calibrate a baseline copy number. In our analysis, the amplicons of the AZFc locus are taken as the on-target region, with the reminder of chrY constituting the antitarget region.

Copy number quantification in CNVkit is performed using a fixed-bin approach in which on- and off-target regions are divided into bin regions of optimal but fixed size (calculated as part of the CNVkit pipeline) for the specific analysis being undertaken. In the CNVkit-ampliconic approach described in this publication, we selected as our on-target regions the same ampliconic loci described by Teitz et al. Copy number was quantified across each fixed bin spanning the AZFc locus, and specific AZFc-CNVs could then be inferred by comparing the resulting copy number predictions to expected patterns of ampliconic copy number change, as for the Teitz-ampliconic approach (Supplemental Table 2).

#### Development of median centering approach for correction of biases in copy number estimates

With sufficient sample size, the peaks of predicted copy number distributions would be expected to be centered at log-fold change values equating to integer fold change values (under the assumption that the targeted regions are either entirely present, entirely absent, or entirely duplicated one or more times in each sample). Further, the modal (i.e. highest) peak of the distributions should lie approximately at a log-fold change value of 0, representing a fold change value of 1 (i.e. no change) compared to the reference.

Following the initial run of the two gr/gr deletion calling methodologies on the high-coverage 1KGP WGS data, it was observed that the resulting copy number estimates of both STS marker regions were slightly offset from the expected integer copy numbers. These results meant that the modal peaks, expected to correspond to the reference ampliconic copy numbers, lay a little to the above or below the reference log-fold change value of 0 (Supplemental Figure 2). The directionality of this effect differed at different levels of sequencing coverage and varied between amplicons, suggests that this divergence may act in a region-/marker-specific manner.

To address this issue, we incorporated a corrective “median centering” adjustment into the two calling approaches to move the modal peak of each distribution to align with a log-fold change of 0. The magnitude of adjustment was calculated by calculating the difference between the sample-wide median log-fold change value and zero, and then subtracting this value from all constituent log-fold change values (Supplemental Figure 2). As the reference copy number of each amplicon is the predominant copy number across samples, the median of each distribution was deemed a suitable approximation for the mode. It should be noted that this approach may not be suitable for markers or regions with greater copy number heterogeneity (i.e. those where no specific copy number predominates across the sampled population), as, in such cases, the median value may not lie in the proximity of the modal peak. This adjustment was integrated into the pipelines of both approaches prior to the generation of the results displayed in the main publication.

#### Collection and sequencing of TECAC cohort

Samples from 4,821 male subjects across 11 contributing centers were collected and sent to Gencove for processing. DNA extraction, low-pass whole-genome sequencing (WGS) at 0.5 depth of coverage, and imputation were performed by Gencove using a proprietary pipeline. WGS data was imputed against the 1000 Genomes reference panel using GLIMPSE2 (Rubinacci et al., 2023; Li & Stephens, 2003) and custom tools. 81 samples were excluded due to an absence of associated phenotype data.

#### STS-PCR and qPCR validation of computational predictions of gr/gr deletions in the TECAC cohort

To evaluate the accuracy of our computational approaches in identifying gr/gr deletions, we identified a validation cohort of individuals for which to use orthogonal PCR-based approaches. From the 3119 samples in the ultra-low coverage TECAC cohort, 79 were selected for this validation cohort based on the availability of sufficient remaining DNA for further wet lab investigation.

We first conducted standard STS-PCR in these samples (Supplemental Figure 2). All samples were tested for the b2/b4 deletion (encompassing the whole AZFc region) using sY254 and sY255 STS markers and for the gr/gr deletions using sY1191 and sY1291. sY14 (mapping to the SRY locus) was used as an internal control to indicate presence of the Y chromosome. Samples from normal fertile men without Y chromosome microdeletions were used as normal controls.

For the two PCR reactions (b2/b4 and gr/gr) a total of 50-100 ng of genomic DNA was used as template in 25 µL reaction mix, 10X amplification buffer, 25mM MgCl2, 2.5mM dNTPs, 10X primers mix and 5U/ul Taq DNA polymerase (Experteam, Venice, Italy). After an initial denaturation step of 5 min, each PCR reaction was carried out at the annealing temperature specific for STS primers (Supplemental Table 3), ended by an elongation step of 7 min and cooled to 4°C. DNA amplicons were assessed on the Agilent 200 TapeStation with D1000 ScreenTape assays (Agilent Technologies, Santa Clare, CA, USA). Exemplar STS-PCR gel images are given in Supplemental Figure 2. For STS-PCR, gr/gr deletions were called on the basis of absence of the sY1291 marker with presence of the sY1191 marker.

We additionally carried out qPCR for more precise quantification of STS target amplicons in a subset of five samples (Supplemental Figure 2). As above, any sample with a significantly decreased level of sY1291 marker, and no significant change in sY1191, was interpreted as carrying a gr/gr deletion. qPCR was performed in a 20 µl final volume containing 20 ng of cDNA, 1X Power SYBR Green PCR Master Mix (Applied Biosystem, Foster City, CA, USA), and a mix of forward and reverse primers (1 mmol/l each; Supplemental Table 3). The sY1191 and sY1291 STS regions were selected for qPCR amplification to allow corroboration of gr/gr deletions. The human 36B4 gene was used for normalization. qPCR was performed on thermocycler StepOne plus (Applied Biosystems, Foster City, CA, USA) and relative quantification was performed using the 2-delta delta Ct (ΔΔCt) method.

#### Ascertainment of UK Biobank TGCT cohort

To generate a suitable set of UK Biobank (UKB) TGCT cases and controls for association analysis, we first extracted the following fields for all participants via the cohort browser utility of the UKB:

- Sex (p31)
- Genetic sex (p22001)
- Cancer code, self-reported (p20001)
- Type of cancer: ICD10 (p40006)
- Type of cancer: ICD9 (p40013)
- Histology of cancer tumour (p40011)
- Behaviour of cancer tumour (p40012)

Male participants were identified by filtering to retain only those with concordant values of “Male” for both the sex (p31) and genetic sex (p22001) fields.

These white male participants were divided into case and control cohorts. Individuals with a history of malignant testicular cancer were identified using any of the following ICD10 or ICD9 codes was present in their registry-linked “Type of cancer” fields AND where the “Behaviour of cancer tumour” field indicated confirmed malignancy of the tumour:

| **ICD10** | **ICD9** |
| --- | --- |
| C62.0 Undescended testis | 1869 Malignant neoplasm of testis, other and unspecified |
| C62.1 Descended testis |  |
| C62.9 Testis, unspecified |  |

Specific instances of TGCT among the broader testicular cancer cohort were identified using the “Histology of cancer tumour” field. Individuals with any of the below annotations were assigned to both an overall TGCT case cohort and the respective histological subcohort of either “seminoma” or “non-seminoma/mixed”; histological annotations of “spermatocytic seminoma” were included in the overall TGCT cohort but not in either subgroup due to its distinct histopathological features. Similarly, any individuals with recorded instances of both seminoma and non-seminoma/mixed cancers were included only in the overall TGCT cohort.

| **Seminoma** | **Non-seminoma/Mixed** | **Other** |
| --- | --- | --- |
| Seminoma, NOS | Teratoma, malignant, NOS | Spermatocytic seminoma |
| Seminoma, anaplastic | Mixed germ cell tumour |  |
|  | Malignant teratoma, undifferentiated |  |
|  | Malignant teratoma, intermediate |  |
|  | Germinoma |  |
|  | Embryonal carcinoma, NOS |  |
|  | Teratocarcinoma |  |
|  | Malignant teratoma, trophoblastic |  |
|  | Dysgerminoma |  |
|  | Yolk sac tumour |  |
|  | Choriocarcinoma combined with other germ cell elements |  |
|  | Choriocarcinoma |  |

We additionally included any individuals with an instance of self-reported “testicular cancer” in our case cohort, with the exception of individuals who had registry-linked records of non-TGCT testicular cancer. All samples not assigned to the case cohort were automatically assigned to the control cohort.

#### Principal component analysis for identification of samples of European ancestry

To filter for individuals of high-confidence European ancestry, a pooled dataset was constructed comprising autosomal SNP array data from a subset of UKB samples (n=222,079) plus the WGS data from the TECAC (n=4728) and 1KGP (n=1194) cohorts. SNPs were filtered to retain only those present in all three datasets and PCA conducted on the filtered dataset using Plink^3^ (Supplemental Figure 3). A random forest model was trained using the Python library scikit-learn to predict the corresponding superpopulation for 70% of the labelled 1KGP samples using the first ten principal components (PCs) as features. This model exhibited >0.98 accuracy in classifying the remaining (30%) unseen 1KGP samples as being of European ancestry.

This model was then applied to the first ten PCs of the UKB and TECAC data and samples predicted by the random forest to be of European ancestry retained for the gr/gr deletion-TGCT association analysis. To account for residual variability in TGCT risk even within European populations, a second round of dataset-specific PCA was conducted within the UKB and TECAC European cohorts using the larger set of SNP markers made available by not pooling the data. The top three PCs of this analysis were then integrated as covariates into the final logistic regressions.

### Supplemental Figures

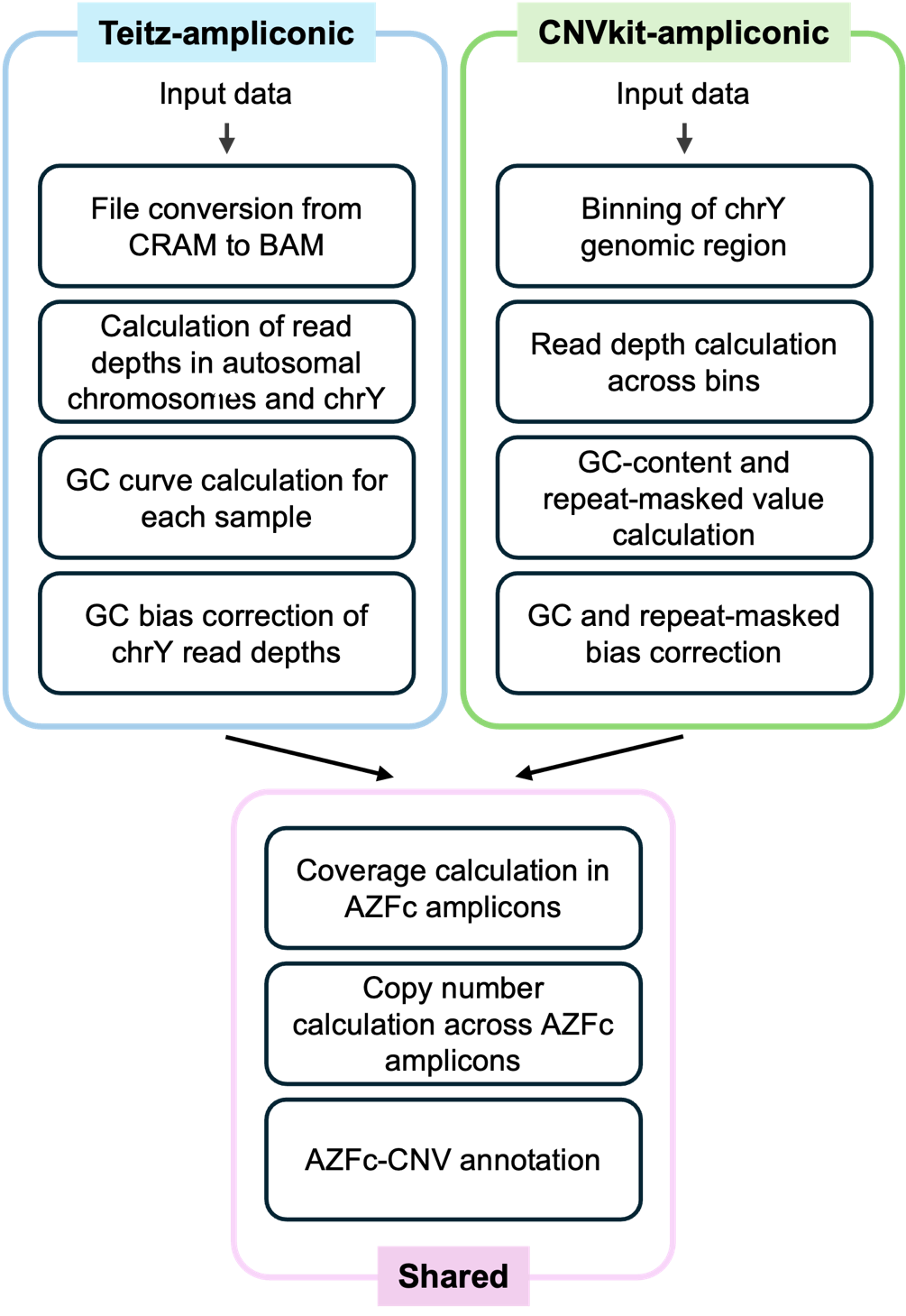

**Supplemental Figure 1. Workflow schematic for two bioinformatics approaches to AZFc-CNV calling.** Shown are the stages of AZFc-CNV calling for the Teitz-ampliconic approach as previously published^1^ and the CNVkit-ampliconic approach described in the main manuscript. The two pipelines differ in the input files required (CRAM vs. BAM), the genomic regions used to establish baseline reference copy number (autosomal chromosomes in Teitz-ampliconic and chrY in CNVkit-ampliconic) and the GC and repeat-masking correction approaches. Coverage calculations and AZFc-CNV inference in each sample are conducted using the published Teitz-ampliconic pipeline for both approaches.

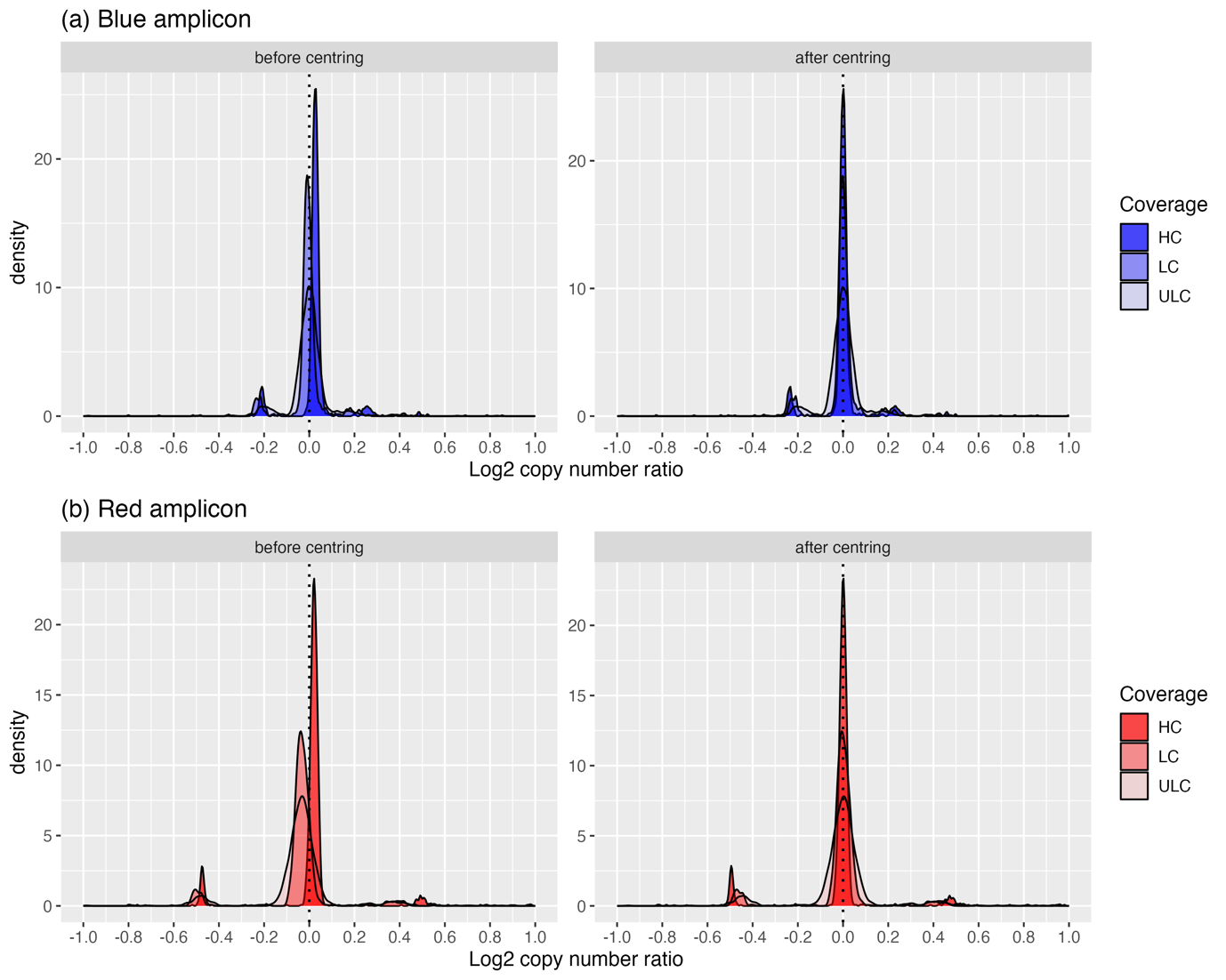

**Supplemental Figure 2.** **Effect of median centering adjustment on predicted amplicon copy number ratio distributions in the Teitz-ampliconic approach for gr/gr deletion detection.** Shown are the distributions of predicted copy number ratios across two exemplar amplicons, (**A**) blue and (**B**) red, used to identify gr/gr deletions as predicted by the Teitz-ampliconic approach in 1194 1KGP samples at high- (HC), low- (LC) and ultra-low- (ULC) coverage (left) before and (right) after integration of a batch median centering adjustment.

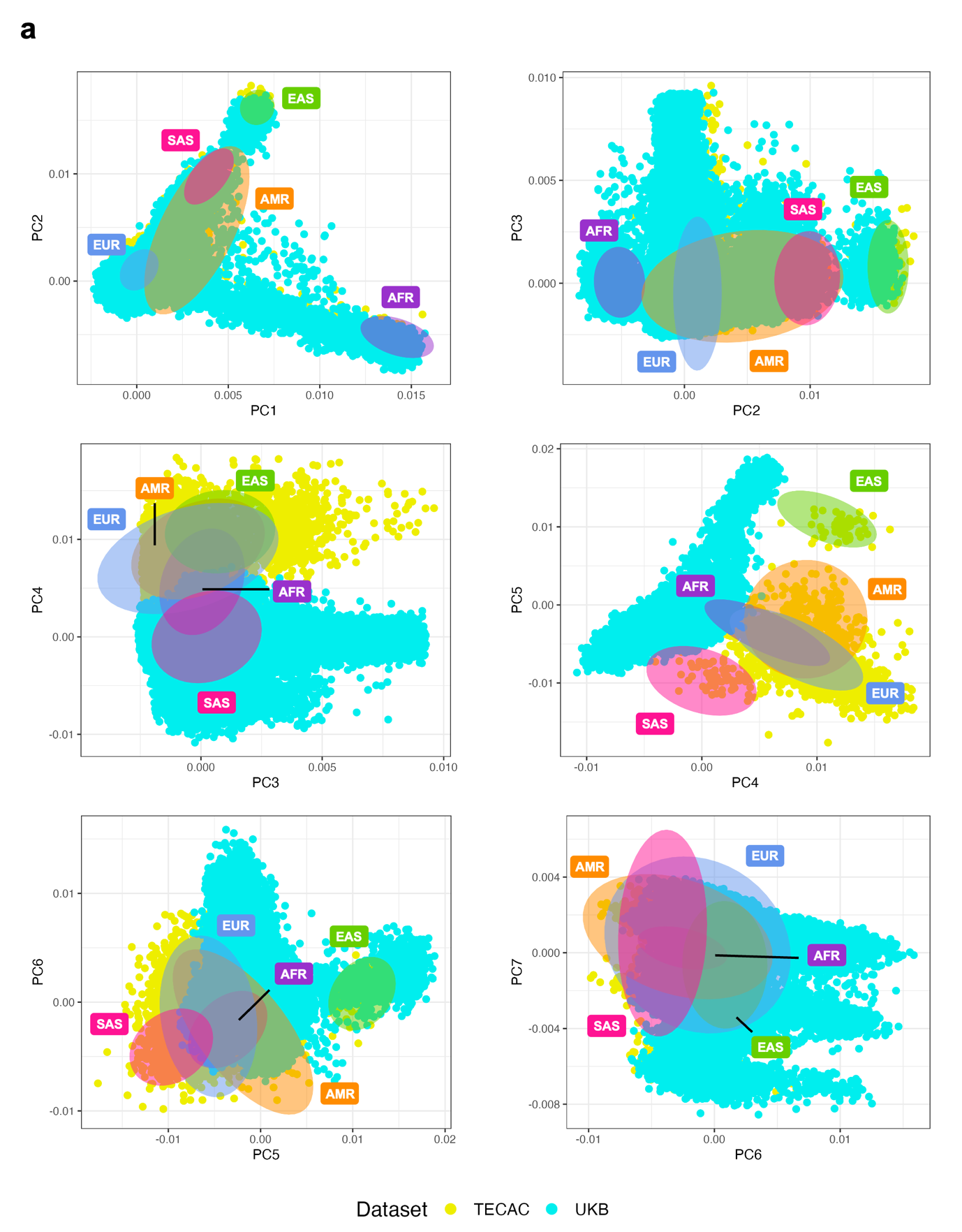

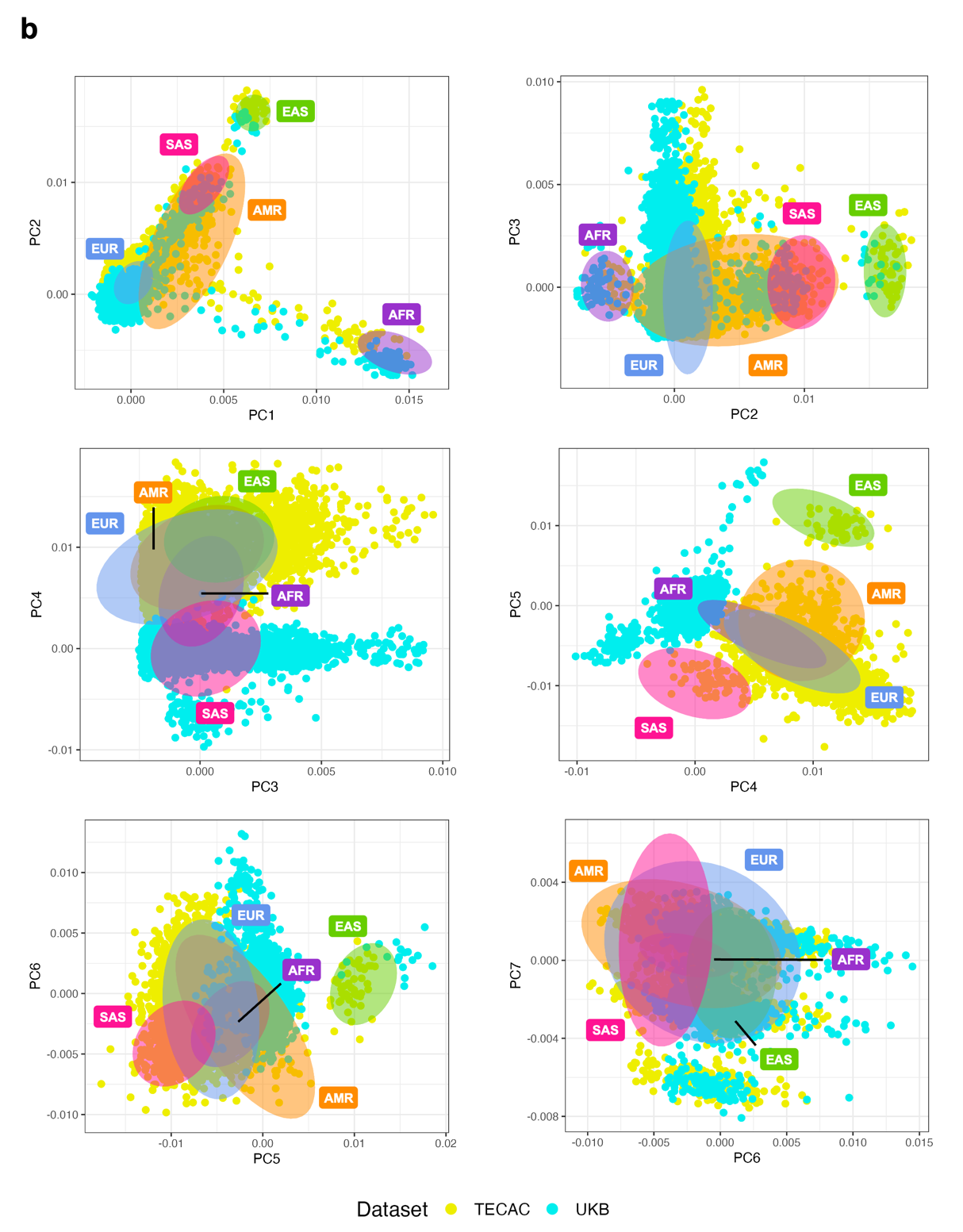
Supplemental Figure 3. Principal component analysis of UKB and TECAC samples. As an initial step to identify individuals of European-like ancestry in our study cohorts, principal component analysis (PCA) was performed on pooled male UKB (n=221,979) and TECAC (n=4728) samples alongside samples from the 1000 Genomes Project (1KGP; n=1194) which had existing ancestry annotations. Shown are the distribution of TECAC and UKB samples across successive pairwise combinations of the first seven principal components (PCs), alongside 1KGP samples stratified by superpopulation (coloured ellipses, individual 1KGP data points not shown). We observe good separation of 1KGP superpopulations across these first seven PCs. The large TECAC and UKB cluster within the European superpopulation across PC1 and PC2 indicates the high prevalence of European ancestry in both datasets. Data are shown for TECAC and each of: (A) the full UK Biobank cohort and, for ease of visualization, (B) a randomly selected subset of 4728 UKB samples (matching the number of TECAC samples). AFR, African; AMR, Admixed American; EAS, East Asian; EUR, European; SAS, South Asian.

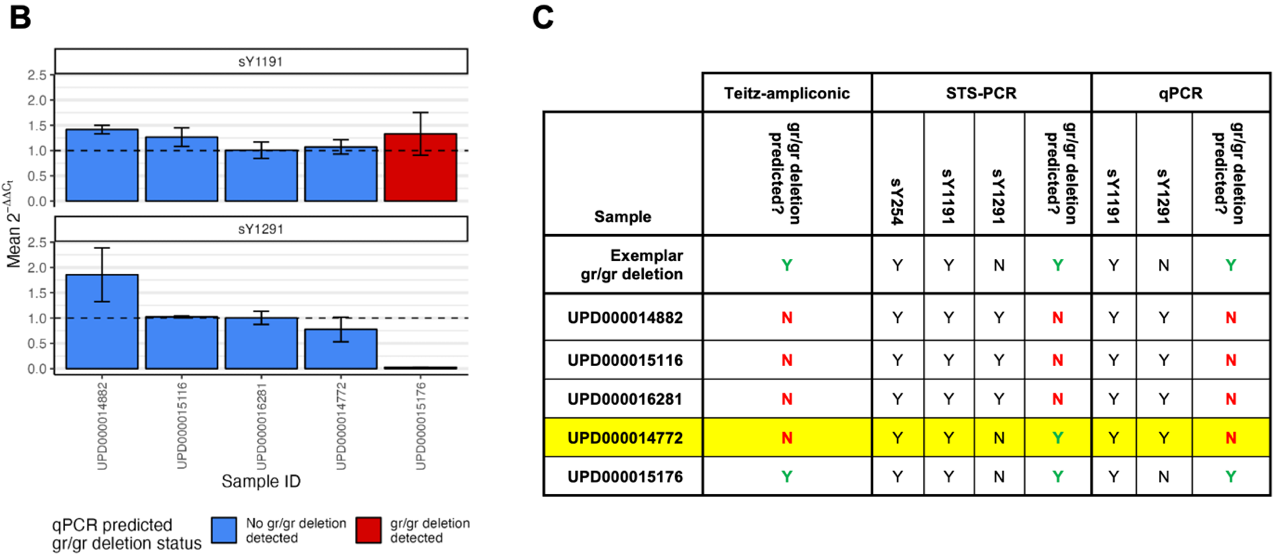

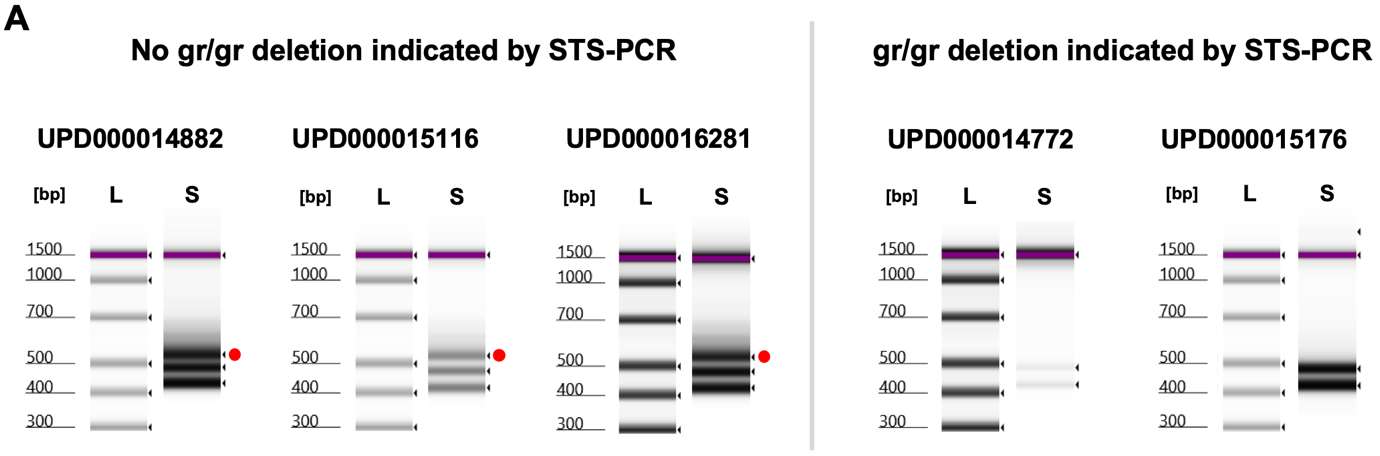

**Supplemental Figure 4. Validation of computational predictions of gr/gr deletion hemizygote status in five male samples using STS-PCR and qPCR.** (**A**) STS-PCR results corroborated computational predictions for the sample predicted to harbor a gr/gr deletion event (UPD000015176) and three of the four samples with predicted to be gr/gr-normal. For one computationally predicted gr/gr-normal sample (UPD000014772), the STS-PCR results were instead indicative of the presence of a gr/gr deletion due to the lack of observed amplification of the sY1291 marker (red dots on left panel). (**B**) More precise real-time quantification of STS marker amplicons using qPCR showed full concordance with computationally predicted gr/gr deletion status across all samples: in contrast with the STS-PCR data, qPCR of UPD000014772 indicated the presence of the sY1291 marker in the sample, albeit with higher variability between replicates, evidenced by larger error bars. This sample was of older age, and DNA concentration as measured by Qubit™ dsDNA BR Assay Kit was indeed lower than that determined by the NanoDrop Spectrophotometer, indicating poor DNA quality of the sample. The horizontal dashed line indicates the threshold for relative quantification below which the samples are likely deleted; (**C**) Summary table of predicted gr/gr deletion status of the five samples via the Teitz-ampliconic computational approach, and via the presence (Y) or absence (N) of diagnostic markers using the wet-lab STS-PCR and qPCR approaches. The sample with discrepant findings between the two PCR-based approaches is highlighted in yellow and the expected indicative marker presence for a typical gr/gr deletion given in the first row. Original gel images were cropped and universally resized to ensure retention of accurate dimensional proportions for each sample. L, ladder lane; S, sample lane.

### Supplemental Tables

| Centre |  | No. samples (pre-QC) | | | No. samples (post-QC) | | |
| --- | --- | --- | --- | --- | --- | --- | --- |
|  | **Country** | **Men with TGCT** | **Men without TGCT** | **Total samples** | **Men with TGCT** | **Men without TGCT** | **Total samples** |
| CNIO | Spain | 135 | 324 | 459 | 75 | 149 | 224 |
| CNIO-CAG | Spain | 465 | 352 | 817 | 217 | 160 | 377 |
| Florence | Italy | 82 | 78 | 160 | 43 | 39 | 82 |
| Groningen | Netherlands | 392 | 69 | 461 | 215 | 49 | 264 |
| Leeds | UK | 746 | 123 | 869 | 378 | 59 | 437 |
| MD Anderson | USA | 604 | 303 | 907 | 214 | 115 | 329 |
| Minnesota | USA | 58 | 0 | 58 | 21 | 0 | 21 |
| Padova | Italy | 79 | 86 | 165 | 43 | 40 | 83 |
| Pennsylvania | USA | 348 | 0 | 348 | 139 | 0 | 139 |
| Princess Margaret | Canada | 332 | 51 | 383 | 197 | 30 | 227 |
| Turin | Italy | 70 | 12 | 82 | 32 | 6 | 38 |
| Other | - | 31 | 0 | 31 | 10 | 0 | 10 |
| All contributing centres | | **3342** | **1398** | **4740** | **1584** | **647** | **2231** |

**Supplemental Table 1. Counts of *WGS* samples in TECAC dataset stratified by contributing centre.** Shown are the number of starting numbers of samples with available WGS data from each of the TECAC contributing centres, and the number remaining after following the filtering steps detailed in Supplemental Table 4.

| AZFc-CNV ID | Established  name for CNV | IR5 | Blue | Green | Red | Gray | Yellow | IR1 | Teal |
| --- | --- | --- | --- | --- | --- | --- | --- | --- | --- |
| 1 | gr/gr deletion | -1 | -1 | -1 | -2 | -1 | -1 | 0 | 0 |
| 2 | gr/gr duplication | 1 | 1 | 1 | 2 | 1 | 1 | 0 | 0 |
| 3 | b2/b3 deletion | -1 | -1 | -2 | -2 | 0 | -1 | -1 | 0 |
| 4 | b2/b3 duplication | 1 | 1 | 2 | 2 | 0 | 1 | 1 | 0 |
| 5 | gr/gr rescue | 0 | 0 | 1 | 0 | -1 | 0 | 1 | 0 |
| 6 | b2/b3 rescue | 0 | 0 | -1 | 0 | 1 | 0 | -1 | 0 |
| 7 | b2/b4 duplication | 2 | 2 | 3 | 4 | 1 | 2 | 1 | 0 |
| 8 | b1/b3 duplication | 0 | 2 | 1 | 2 | 1 | 0 | 1 | 2 |
| 9 | b1/b3 deletion | 0 | -2 | -1 | -2 | -1 | 0 | -1 | -2 |
| 10 | - | 2 | 2 | 2 | 4 | 2 | 2 | 0 | 0 |
| 11 | - | 2 | 2 | 4 | 4 | 0 | 2 | 2 | 0 |
| 12 | - | 4 | 4 | 4 | 8 | 4 | 4 | 0 | 0 |
| 13 | - | 3 | 3 | 6 | 6 | 0 | 3 | 3 | 0 |
| 14 | - | 1 | 1 | 3 | 2 | -1 | 1 | 2 | 0 |

**Supplemental Table 2.** **Ampliconic copy number changes indicative of various classes of AZFc-CNV using the Teitz-ampliconic approach.** Shown are the copy number changes expected to result from each class of NAHR-based AZFc-CNV. Copy number changes observed in sequencing data can be readily mapped back to corresponding classes of AZFc-CNVs. Adapted from the GitHub repository containing the codebase for the work presented in Teitz et al.^1^ (<https://github.com/lsteitz/y-amplicon-evolution>, accessed 7^th^ September 2025).

| **Gene/STS marker** | **Function in analysis** | **Primer direction** | **Primer sequence** |
| --- | --- | --- | --- |
| **sY14**  **(SRY)** | Internal control to show presence of Y-chromosome in STS-PCR samples | **F** | GAATATTCCCGCTCTCCGGA |
|  |  | **R** | GCTGGTGCTCCATTCTTGAG |
| **36B4** | Single-copy gene against which qPCR expression levels are normalized | **F** | CAGCAAGTGGGAAGGTGTAATCC |
|  |  | **R** | CCCATTCTATCATCAACGGGTACAA |
| **sY1191** | STS marker required to be present for gr/gr deletion calling in STS-PCR and qPCR | **F** | CCAGACGTTCTACCCTTTCG |
|  |  | **R** | GAGCCGAGATCCAGTTACCA |
| **sY1291** | STS marker required to be absent for gr/gr deletion calling in STS-PCR and qPCR | **F** | GGGAGAAAAGTTCTGCAACG |
|  |  | **R** | TAAAAGGCAGAACTGCCAGG |
| **sY254** | STS marker used to exclude gross AZFc deletions in STS-PCR | **F** | GGGTGTTACCAGAAGGCAAA |
|  |  | **R** | GAACCGTATCTACCAAAGCAGC |
| **sY255** | STS marker used to exclude gross AZFc deletions in STS-PCR | **F** | GTTACAGGATTCGGCGTGAT |
|  |  | **R** | CTCGTCATGTGCAGCCAC |

**Supplemental Table 3.** **Forward and reverse primer sequences for amplification of STS markers used for gr/gr deletion characterization in STS-PCR and qPCR approaches.** F, forward; R, reverse.

| **Exclusion criterion** | **TECAC** | | **UKB** | |
| --- | --- | --- | --- | --- |
|  | **Samples excluded** | **Samples remaining**  **(with/without TGCT)** | **Samples excluded** | **Samples remaining**  **(with/without TGCT)** |
| Total samples (pre-QC) | - | 4740  (3342/1398) | - | 221,979 |
| Failed to run Teitz-ampliconic scripts, or single-read coverage <0.05 | 12 | 4728  (3336/1392) | 0 | 221,979 |
| Non-European predicted ancestry | 615 | 4113  (2862/1251) | 14,144 | 207,835  (821/207,014) |
| Samples with copy number changes at single amplicons^a^ | 1139 | 2974  (2091/883) | 11,760 | 203,985  (810/203,175) |
| Not gr/gr deletion-positive or AZFc wild-type | 743 | 2231 | 7910 | 196,075 |
| **Total men with TGCT^b^** | 1584  (370 seminoma; 318 non-seminoma/mixed; 1 spermatocytic seminoma; 895 lacking histology data) | | 785  (432 seminoma; 211 non-seminoma/mixed; 6 spermatocytic seminoma; 6 multiple histological types; 130 lacking histology data) | |
| **Total men without TGCT** | 647 | | 195,290 | |

**Supplemental Table 4.** **Counts of TECAC and UKB samples filtered by chrY-specific and ancestry-related exclusion criteria prior to association analysis.** Shown are the stepwise exclusion criteria implemented following running of the Teitz-ampliconic pipeline on eligible samples (all samples with phenotype data for TECAC; all samples with available SNP array data for UKB).

^a^ Samples with predictions of single-amplicon copy number changes using the Teitz-ampliconic approach were excluded based on a high likelihood of false-positive calls, as per guidance in the original publication^1^.

^b^ Histology data was only available for a subset of 689 men with TGCT in the TECAC dataset after QC filtering. These numbers may increase as more clinical data becomes available. Two UKB samples and one TECAC sample from men with TGCT had a histology annotation of “spermatocytic seminoma” and were deemed unsuitable for inclusion in either the seminoma or non-seminoma/mixed subgroups (see Supplemental Methods).

| **TGCT histology** | **Dataset** | **# gr/gr deletion hemizygotes/total (%)** | | | **aOR**  **(95% CI)** | ***p*-value** |  | |
| --- | --- | --- | --- | --- | --- | --- | --- | --- |
|  |  | **Men with TGCT** | **Men without TGCT** | **All samples** |  |  | **Cochran’s Q (*p*-value)** | **I^2^**  **(95% CI)** |
| **All TGCTs** | UKB | 11/785  (1.40%) | 2932/195,290  (1.50%) | 2943/196,075  (1.50%) | 0.98  (0.54-1.75) | 0.93 | 1.45  (0.23) | 30.95  (0-91.2) |
|  | TECAC | 38/1584  (2.40%) | 12/647  (1.85%) | 50/2231  (2.24%) | 1.69  (0.86-3.33) | 0.12 |  |  |
|  | Meta-analysis  (RE) | 49/2369  (2.07%) | 2944/195,937  (1.50%) | 2993/198,306  (1.51%) | **1.24**  **(0.74-2.07)** | **0.42** |  |  |
|  | Meta-analysis  (FE) | 49/2369  (2.07%) | 2944/195,937  (1.50%) | 2993/198,306  (1.51%) | **1.23**  **(0.79-1.92)** | **0.36** |  |  |
| **Seminoma** | UKB | 6/432  (1.39%) | 2932/195,290  (1.50%) | 2938/195,722 (1.50%) | 1.00  (0.46-2.17) | 1.00 | 0.78  (0.38) | 0  (0-88.6) |
|  | TECAC | 9/370  (2.43%) | 12/647  (1.85%) | 21/1017 (2.06%) | 1.70  (0.70-4.09) | 0.25 |  |  |
|  | Meta-analysis  (RE) | 15/802  (1.87%) | 2944/195,937  (1.50%) | 2959/196,739  (1.50%) | **1.26**  **(0.71-2.25)** | **0.43** |  |  |
|  | Meta-analysis  (FE) | 15/802  (1.87%) | 2944/195,937  (1.50%) | 2959/196,739  (1.50%) | **1.26**  **(0.71-2.25)** | **0.43** |  |  |
| **Non-seminoma/**  **mixed** | UKB | 2/211  (0.95%) | 2932/195,290  (1.50%) | 2934/195,501 (1.50%) | 0.78  (0.23-2.71) | 0.69 | 0.88  (0.35) | 0  (0-89.0) |
|  | TECAC | 8/318  (2.52%) | 12/647  (1.85%) | 20/965  (2.07%) | 1.64  (0.66-4.01) | 0.30 |  |  |
|  | Meta-analysis  (RE) | 10/529  (1.89%) | 2944/195,937  (1.50%) | 2954/196,466  (1.50%) | **1.26**  **(0.61-2.61)** | **0.53** |  |  |
|  | Meta-analysis  (FE) | 10/529  (1.89%) | 2944/195,937  (1.50%) | 2954/196,466  (1.50%) | **1.26**  **(0.61-2.61)** | **0.53** |  |  |

**Supplemental Table 5.** **Estimated effect sizes for association between gr/gr deletions and TGCT, including under fixed-effect meta-analysis.** Shown are the estimated effect sizes in constituent datasets and upon random-effects inverse variance-weighted meta-analysis. Further to these data presented in Table 2, the results of a fixed-effect inverse variance-weighted meta-analysis are additionally presented here. For the histological subgroup analyses, the fixed-effect and random-effects estimates are equivalent, as the I^2^ metric equaled 0, thus providing no extra relative weighting to the studies. aOR, adjusted odds ratio; FE, fixed-effect; RE, random-effects.

| **Subject group** | **gr/gr deletion** | | **OR (95% CI)** | **aOR^a^ (95% CI)** | **aOR^b^ (95% CI)** |
| --- | --- | --- | --- | --- | --- |
|  | ***n*** | **%** |  |  |  |
| Unaffected men^c^  (*n* = 2599) | 33 | 1.3 | 1.0 | 1.0 | 1.0 |
| TGCT cases  (*n* = 1842) | 42 | 2.3 | 1.8 (1.1-2.9) | 2.1 (1.3-3.6) | 2.1 (1.3-3.5) |
| Positive family history^d^  (*n* = 431) | 13 | 3.0 | 2.4 (1.3-4.6) | 3.2 (1.5-6.7) | 2.9 (1.2-6.9) |
| Negative family history^d^  (*n* = 1376) | 28 | 2.0 | 1.6 (0.97-2.7) | 1.9 (1.1-3.3) | 1.9 (1.1-3.4) |
| Paternal lineage^e^  (*n* = 345) | 6 | 1.7 | 1.4 (0.57-3.3) | 1.6 (0.54-5.0) | 1.7 (0.50-5.9) |
| Maternal lineage^e^  (*n* = 80) | 7 | 8.8 | 7.5 (3.2-17) | 9.8 (3.5-27) | 9.4 (3.0-29) |
| Seminoma^f^  (*n* = 827) | 27 | 3.3 | 2.6 (1.6-4.4) | 3.0 (1.6-5.4) | 2.8 (1.4-5.2) |
| Positive family history  (*n* = 146) | 6 | 4.1 | 3.3 (1.4-8.1) | 3.3 (1.2-9.4) | 2.3 (0.51-9.9) |
| Negative family history  (*n* = 660) | 20 | 3.0 | 2.4 (1.4-4.3) | 2.7 (1.4-5.2) | 2.8 (1.5-5.5) |
| Nonseminoma^f^  (*n* = 806) | 13 | 1.6 | 1.3 (0.67-2.4) | 1.5 (0.72-3.0) | 1.7 (0.79-3.5) |
| Positive family history  (*n* = 145) | 5 | 3.5 | 2.8 (1.1-7.2) | 4.9 (1.7-14) | 4.0 (1.1-14) |
| Negative family history  (*n* = 653) | 8 | 1.2 | 0.96 (0.44-2.1) | 1.3 (0.55-3.0) | 1.4 (0.60-3.3) |

##### **Supplemental Table 6. Stratified estimated effect sizes of the association between gr/gr deletions and TGCT in Nathanson et al. (2005).** Data taken directly from the source publication^4^.

^a^ Adjusted for study center (Philadelphia, western Washington State, other North America, Hungary, United Kingdom and other Europe or Australia).

^b^ Includes only those study centers that contributed both patients with TGCT and unaffected men (Hungary, Philadelphia, United Kingdom and western Washington State) and is adjusted for those study centers.

^c^ Reference group for all comparisons.

^d^ Information on family history of TGCT was not available for 23 cases (17 seminoma and 6 nonseminoma), including 1 gr/gr deletion carrier. In addition, 12 cases (4 seminoma, 2 nonseminoma, and 6 of unknown tumor type) with an affected [monozygotic] twin were excluded from analyses of family history.

^e^ Information on six TGCT cases with a family history (one from the ITCLC familial studies and five from the case-control studies) was insufficient to determine the type of Y-chromosome transmission.

^f^ Information on tumor type was not available for 209 TGCT cases, including 2 gr/gr deletion carriers.

| **Subject group** | **Prevalence of gr/gr deletion** | |
| --- | --- | --- |
|  | ***n*** | **%** |
| Probands of multiple-case families^a^: |  |  |
| ITCLC family probands | 13/396 | 3.3 |
| Case-series family probands^b^ | 0/35 | 0 |
| Total | 13/431 | 3.0 |
| TGCT sporadic case series^c^: |  |  |
| London | 12/419 | 2.9 |
| Leeds (United Kingdom) | 2/263 | 0.8 |
| Rotterdam | 4/311 | 1.3 |
| Toronto | 2/14 | 14.3 |
| Hungary | 0/18 | 0 |
| Other (Germany, Ireland and Russia)^d^ | 0/8 | 0 |
| Total | 20/1,033 | 1.9 |
| TGCT sporadic cases from case-control series: |  |  |
| Philadelphia | 3/99 | 3.0 |
| Washington State^e^ | 5/167 | 3.0 |
| Total | 8/266 | 3.0 |
| Affected individuals: |  |  |
| With solitary TGCT^f^ | 0/17 | 0 |
| With bilateral TGCT^g^ | 0/61 | 0 |
| Total | 0/78 | 0 |
| Unaffected males: |  |  |
| U.K. control series I | 3/135 | 2.2 |
| U.K. control series II | 1/514 | 0.2 |
| U.K. control series III | 1/225 | 0.4 |
| U.K. control series IV | 7/400 | 1.7 |
| Philadelphia | 5/518 | 0.9 |
| Washington State | 8/435 | 1.6 |
| Hungary | 8/394 | 2.0 |
| Total | 33/2,599 | 1.3 |

**Supplemental Table 7.** **Prevalence of gr/gr deletions among cases and controls in Nathanson et al. (2005).** Data taken directly from source publication^4^.

^a^ Excludes one member from each of 12 [monozygotic] twin pairs.

^b^ Nine of the case-series probands were part of the ITCLC and are included in table 2 [in source publication].

^c^ Individuals with solitary or bilateral TGCT and no family history of TGCT

^d^ These sites are contributors to the ITCLC.

^e^ Does not include 23 individuals with unknown family history.

^f^ Patients ascertained because of family history of [undescended testes].

^g^ Patients ascertained as bilateral cases with no family history of TGCT.
